# Thyroid Cancer Risk Prediction from Multimodal Datasets Using Large Language Model

**DOI:** 10.64898/2026.03.05.26347766

**Authors:** Paramita Ray

## Abstract

Thyroid carcinoma is one of the most prevalent endocrine malignancies worldwide, and accurate preoperative differentiation between benign and malignant thyroid nodules remains clinically challenging. Diagnostic methods that medical practitioners use at present depend on their personal judgment to evaluate both imaging results and separate clinical tests, which creates inconsistency that leads to incorrect medical evaluations. The combination of radiological imaging with clinical information systems enables healthcare providers to enhance their capacity to make reliable predictions about patient outcomes while improving their decision-making abilities. The study introduces a deep learning framework that utilizes multiple data sources by combining magnetic resonance imaging (MRI) data with clinical text to predict thyroid cancer. The system uses a Vision Transformer (ViT) to obtain advanced MRI scan features, while a domain-adapted language model processes clinical documents that contain patient medical history and symptoms and laboratory results. The cross-modal attention system enables the system to merge imaging data with textual information from different sources, which helps to identify how the two types of data are interconnected. The system uses a classification layer to classify the fused features, which allows it to determine the probability of cancerous tumors.

The experimental results show that the proposed multimodal system achieves better results than the unimodal base systems because it has higher accuracy, sensitivity, specificity, and AUC values, which help medical personnel to make better preoperative decisions.

## I. Introduction

### A. Motivation of Work

Accurate diagnosis of thyroid cancer is challenging because they depend on imaging tests which require personal judgment, and its assessment methods lack complete patient information. The MRI scans deliver structural data while the clinical narratives provide essential background information about patient medical history and their test results. The diagnostic process becomes less reliable because healthcare providers tend to examine distinct data elements from various sources instead of evaluating them together.

Recent developments in deep learning technologies, together with large language models (LLMs), create new possibilities for medical data analysis that combine multiple data types to enhance predictive capabilities. This work develops a multimodal framework that combines MRI images with clinical text through cross-modal learning to create more precise thyroid cancer diagnostic support for clinical decision-making. The major contributions of this article can be summed up as follows:

1. An innovative multimodal framework has been developed by integrating deep learning architecture to combining MRI imaging data and clinical text to provide more accurate and reliable thyroid cancer prediction.
2. A cross-model attention module effectively captures the complex relationships between radiological features and clinical representations.
3. The proposed method provides a scalable and highly secure preoperative decision-support framework for detecting early stages of thyroid cancer.

The remainder of the paper is organized as follows. In Section II, we have presented some literature justifying the relevance of using the proposed method, and Section III details the proposed methodology. Section IV contains results, and Section V presents our conclusions.

## II. Related Work

Deep learning has brought major changes to medical image analysis, which specifically benefits oncological diagnostic work. The research shows that Convolutional Neural Networks (CNNs) can effectively detect and classify tumors through their application across different imaging methods [1]. The researchers developed Vision Transformers (ViTs) to solve medical image cancer detection challenges by using their advanced capability to understand entire image contexts [2]. The research studies have used deep learning methods to analyze ultrasound images in thyroid cancer research, which successfully distinguishes between benign and malignant nodules while achieving better results than standard machine learning techniques [3], [4]. The current methods focus on using one type of imaging data because they do not consider additional medical details that could provide useful information. Parallel advancements in Natural Language Processing (NLP) have enabled effective extraction of structured knowledge from unstructured clinical narratives. Transformer-based architectures such as BERT [27] and its clinical adaptations (e.g., ClinicalBERT, BioBERT) have been widely used for medical text classification and risk prediction tasks [5], [6]. More recently, Large Language Models (LLMs) have demonstrated enhanced contextual reasoning and knowledge representation capabilities in biomedical applications [7]. Multimodal learning approaches integrating imaging and clinical data have gained increasing attention for improving diagnostic accuracy and robustness [8,9]. Vision–language models such as CLIP and BLIP enable cross-modal representation learning and alignment between visual and textual features [10]. Despite these advancements, the integration of MRI imaging and clinical text using LLM-driven cross-modal attention mechanisms for thyroid cancer prediction remains underexplored. This gap motivates the development of the proposed multimodal framework.

### A. Research Gap Analysis

Existing deep learning approaches for thyroid cancer prediction predominantly focus on single-modality imaging data, particularly ultrasound-based classification using convolutional neural networks (CNNs). The models show good performance results, but their design restricts them from using multiple types of medical data, which doctors need to evaluate cancerous tumors correctly. The design of CNN-based architectures enables them to capture local spatial features, yet their performance in modeling global contextual dependencies of complex MRI scans remains inadequate.

The application of transformer-based architectures, which include Vision Transformers (ViTs) [11, 13], shows better performance in modeling long-distance spatial patterns, yet their use in analyzing thyroid MRI scans remains underexplored. The transformer-based language models ClinicalBERT [21, 23] and Large Language Models (LLMs) demonstrate effective performance at extracting semantic information from unstructured clinical narratives. Existing multimodal frameworks use basic feature concatenation and late-fusion methods, which do not succeed in capturing advanced cross-modal interaction patterns.

In our proposed work, a multimodal deep learning framework is designed for thyroid cancer prediction that integrates MRI images and clinical text. A Vision Transformer (ViT) [12, 13] extracts spatial features from MRI scans, while Bio-ClinicalBERT encodes contextual information from clinical narratives. The extracted visual and textual representations are fused using a cross-modal attention mechanism to capture interactions between imaging and clinical features. The combined representation is then passed to a classification layer to predict benign or malignant outcomes, which enhances diagnostic accuracy and reliability.

### B. Vision Transformer Model (ViT) for Image Analysis

A vision transfer model [15] uses a pre-trained deep learning architecture (figure-1) to analyze images and transfer learned visual features to a new task. These models use Convolutional Neural Networks (CNNs), which include ResNet and VGG16, as their main technology because these networks were initially developed using the ImageNet dataset. The general structure contains the following components:

**Fig. 1:**
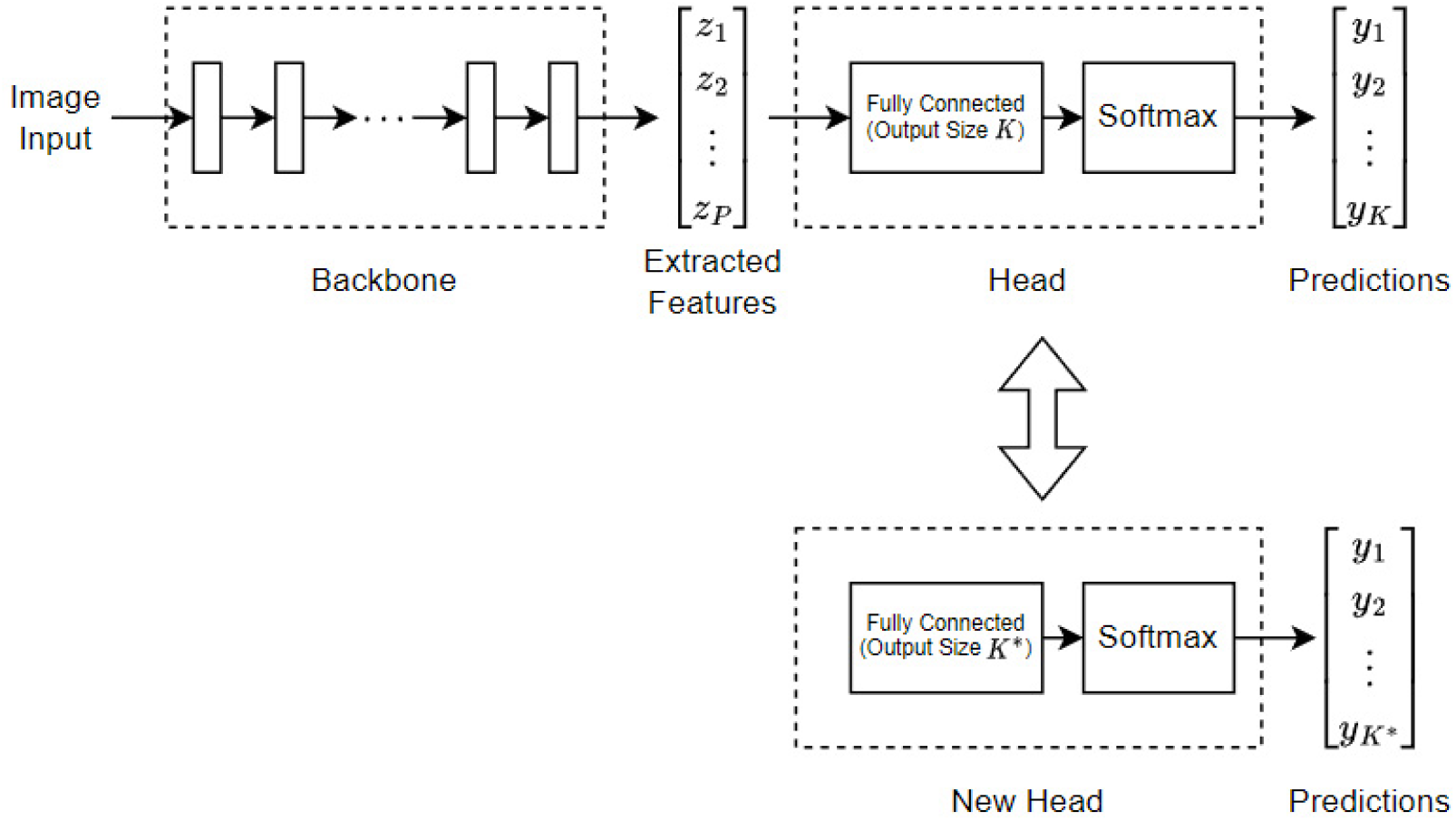
Image analysis using ViT model [25]

### C. Image Representation

An image is represented as a matrix: *I* ∈ *R*^*H×W ×C*^ where H = image height, W = image width, and C = number of channels.

### D. Convolution Operation

The convolution layer extracts visual features from the image.

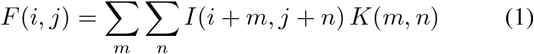

Where: I = input image, K = convolution kernel/filter, F = feature map [17]. This operation detects patterns such as edges, textures, and shapes. Non-linear functions *f* (*x*) = max(0, *x*) help the network learn complex patterns.

### E. Fully Connected Layer

After passing through multiple convolutional [16,18] and pooling layers, the extracted features reach the fully connected layers [19,20], which perform their classification and prediction tasks. The fully connected layer transformation process executes through the following mathematical expression.

z = Wx + b where W denotes the weight matrix, x represents the input feature vector and b stands for the bias term. The softmax function calculates output probabilities for classification tasks. 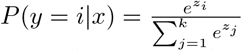 which converts the output scores into normalized probabilities across k classes. During training, the model parameters are optimized by minimizing the cross-entropy loss function. 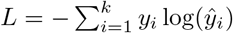 where *y*_*i*_ represents the true label and *ŷ*_*i*_ is the predicted probability. By leveraging transfer learning, vision transfer models significantly reduce training time and improve accuracy, making them highly effective for applications such as medical image diagnosis, object detection, remote sensing, and automated visual inspection.

### F. LLMs for Clinical Text data Analysis

Different large language models (LLMs) are used for text extraction and classification tasks. In this project, clinical text obtained from prescriptions and medical reports is utilized for analysis. Since clinical documents contain specialized medical terminology and contextual information, a domain-specific language model is required for effective feature extraction. Therefore, the BioClinicalBERT [21,22] architecture is employed to extract meaningful representations from the clinical text. BioClinicalBERT is a transformer-based model pre-trained on large-scale clinical datasets, enabling it to capture complex relationships and medical semantics present in electronic health records and clinical narratives.

#### 1) BioClinicalBERT Architecture

BioClinicalBERT is a language model that specializes in processing clinical narratives and biomedical texts within its specific domain. The model uses the BERT architecture as its foundation and undergoes additional training using extensive clinical databases, which include [24] the MIMIC-III clinical database. The model captures complex contextual relationships that exist in electronic health records and medical notes, and clinical documentation. The architecture implements a transformer encoder design, which consists of multiple self-attention layers and feed-forward neural network components. BioClinicalBERT uses the same BERT-base configuration, which includes 12 transformer encoder layers, 12 attention heads, and a hidden representation dimension of 768. The WordPiece tokenizer first tokenizes the input clinical text, which serves as the initial step. The system creates a token representation by merging three types of embeddings, which include token embeddings, positional embeddings, and segment embeddings. The input representation is expressed as

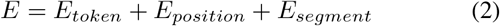

where *E* denotes the final embedding vector used as input to the transformer encoder. Within each transformer layer, contextual relationships between tokens are learned using the self-attention mechanism defined as

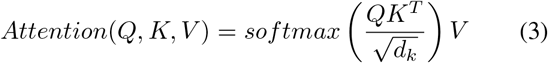

## III. Proposed Methodology

The proposed research presents a multimodal deep learning framework that uses MRI imaging data together with clinical textual information to predict thyroid cancer risk. Medical images show spatial and structural details of thyroid nodules, while clinical narratives provide contextual details about symptom and diagnosis records and treatment history. The system uses both modalities to achieve better diagnostic results, which also enhances decision-making support. The complete system design includes four primary elements, which include data processing, data analysis, and multimodal data integration, and system evaluation. The framework workflow diagram appears in Figure 2.

**Fig. 2:**
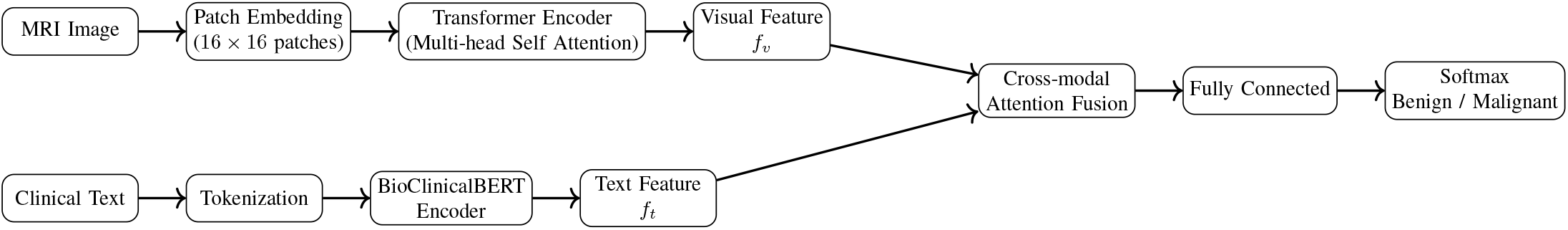
Architecture of the proposed multimodal framework integrating MRI images and clinical text using Vision Transformer and BioClinicalBERT.

### A. MRI Image Feature Extraction using Vision Transformer

MRI images are first preprocessed through resizing, normalization, and noise removal. Let the input MRI image be represented as

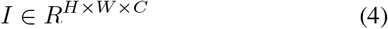

where *H, W*, and *C* denote the height, width, and number of channels of the image.

The Vision Transformer (ViT) divides the image into fixed-size patches of dimension *P* × *P*. Each patch is flattened into a vector representation

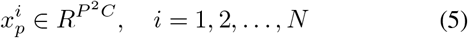

where 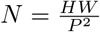 denotes the number of patches.

The patch embeddings are combined with positional encoding to preserve spatial information

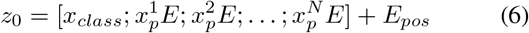

where *E* is the linear projection matrix, and *E*_*pos*_ represents positional embeddings.

The transformer encoder uses a multi-head self-attention mechanism defined as

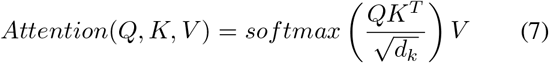

where *Q, K*, and *V* represent query, key, and value matrices, and *d*_*k*_ denotes the dimension of key vectors.

The output of the ViT encoder produces a visual feature vector

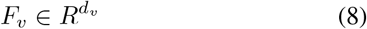

which captures global spatial relationships within the MRI images.

### B. Clinical Text Feature Extraction using BioClinicalBERT

Clinical text obtained from prescriptions and medical reports is processed using BioClinicalBERT. The clinical document is represented as a sequence of tokens

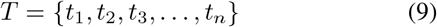

Each token embedding is formed by combining token, positional, and segment embeddings

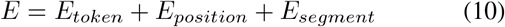

The contextual representation is generated through transformer layers

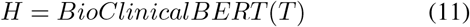

The final textual feature vector is extracted from the special [CLS] token

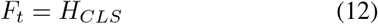

where 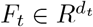 represents the contextual embedding of the clinical text.

### C. Cross-Modal Feature Fusion

To integrate visual and textual representations, a cross-modal attention mechanism is applied. The fusion process allows the model to learn interactions between MRI image features and clinical textual information.

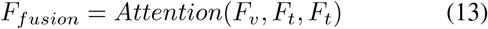

Alternatively, the multimodal feature vector can be represented through concatenation

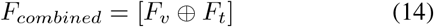

where ⊕ denotes the concatenation operation.

### D. Classification Layer

The fused feature representation is passed to a fully connected layer to perform classification

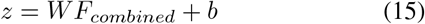

The final prediction is obtained using the softmax function

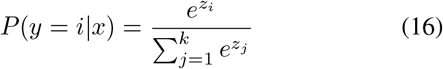

where *k* = 2 corresponds to the benign and malignant classes.

The model is optimized using the cross-entropy loss function

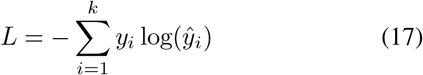

where *y*_*i*_ represents the true label and *ŷ*_*i*_ denotes the predicted probability.

## IV. RESULTS & ANALYSIS

### Data Collection

The collected data set (kaggle public data set (https://www.kaggle.com/datasets/ankushpanday1/thyroid-cancer-risk-prediction-dataset)) includes information on thyroid ultrasound images, patient symptom reports, clinicopathologic features, and demographic characteristics. Every patient in the data set was monitored for a minimum of ten years during the course of the 15-year data collection period.

#### A. Demographic Features Extraction

Demographic features include patient age, gender, smoking habit, history of smoking habits, history of radiotherapy, etc. After analyze the distribution of age with the associated risk of thyroid pathology (Fig-3), it has been observed that risk increases with age.

**Fig. 3:**
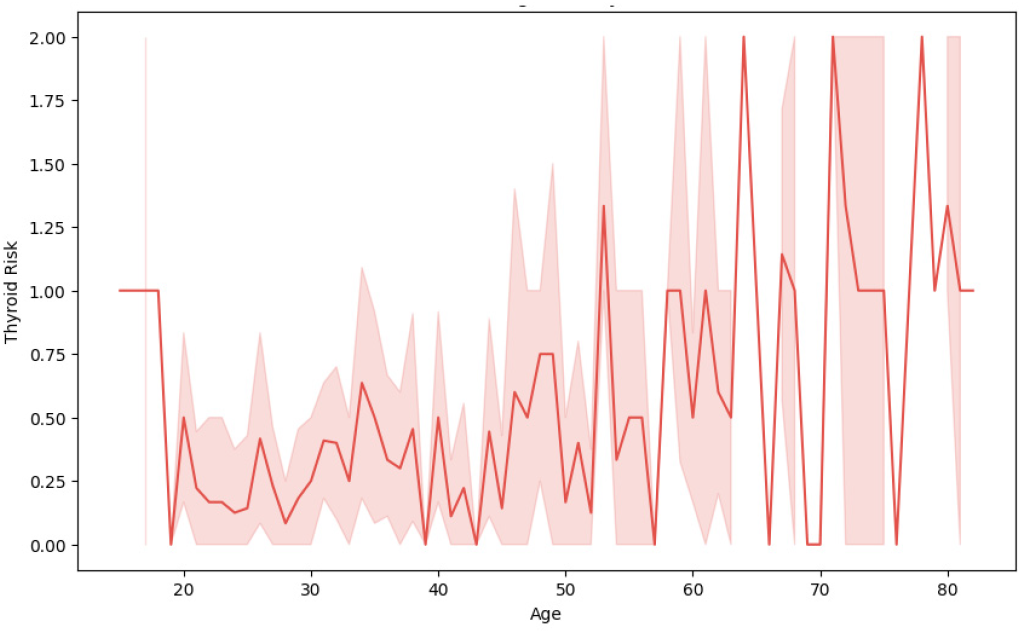
Age distribution based on risk

By calculating (Table-I) and visualizing (Fig-4) the risk ratios of thyroid cancer stages (I, II, III, IV A, IV B) across different age groups, we can observe how the risk of developing each stage of thyroid cancer changes with age. This analysis provides valuable insights into age-related risk factors associated with the progression of thyroid cancer, aiding in the better understanding and management of the disease. The risk ratio for developing Stage 1 thyroid cancer in the 30-40 age group is 0.40, meaning this group has a lower risk compared to other age groups. There are no cases of Stage III thyroid cancer in the sample dataset. The highest risk ratio is in the 60+ age group with a value of 1.60, indicating a significantly higher risk of developing Stage IV thyroid cancer as age increases. The risk tends to increase with age, especially for Stage-IV thyroid cancer, highlighting the importance of age as a risk factor in the progression of thyroid cancer.

**TABLE I:** Risk Ratios of Thyroid Cancer Stages by Age Group.

| Age Group | Stage I | Stage II | Stage III | Stage IV-A | Stage IV-B |
| --- | --- | --- | --- | --- | --- |
| 30 | 0.00 | 0.00 | 0.00 | 0.00 | 0.00 |
| 30-40 | 0.60 | 0.00 | 0.00 | 0.60 | 0.00 |
| 40-50 | 0.00 | 0.00 | 0.00 | 0.60 | 0.00 |
| 50-60 | 0.00 | 0.40 | 0.00 | 0.60 | 0.00 |
| 60+ | 0.00 | 0.00 | 0.60 | 1.60 | 0.40 |

**Fig. 4:**
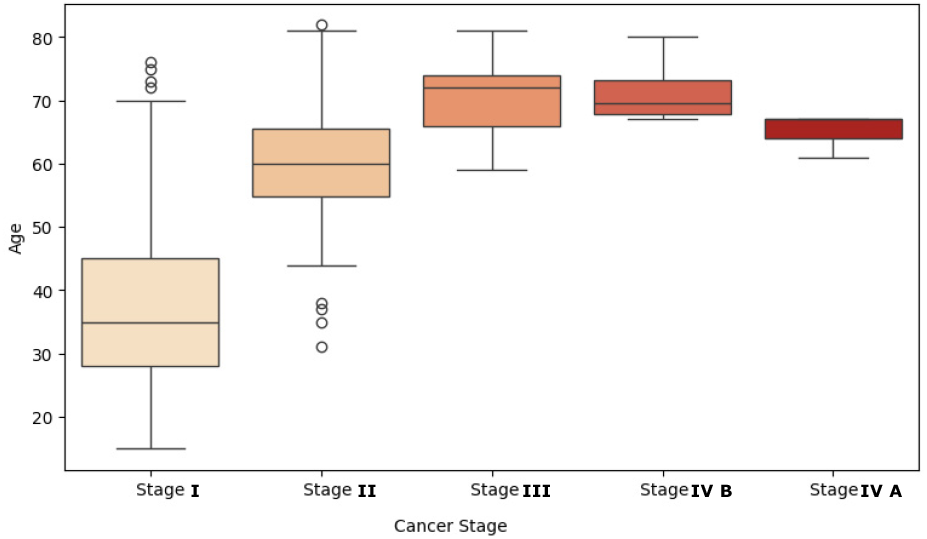
Age distribution based on different stages of Cancer

The TNM system (Tumour, Node, and Metastasis) is typically used for thyroid cancer staging. The main tumor’s size and extent (T), the involvement of nearby lymph nodes (N), and the existence of distant metastases (M) are all taken into consideration by this method. From clinical report, we have got the different size of primary tumors T1 (Tumor ≤ 2 cm), T2 (Tumor *>*2cm but ≤ 4 cm), T3 (Tumor *>* 4) T4(Tumor of any size with extension beyond the thyroid capsule), T4a (Extension into nearby structures such as larynx, trachea, esophagus, or recurrent laryngeal nerve), and T4b (Extension towards the spine or major blood vessels.) Particularly for differentiated thyroid tumours like papillary and follicular thyroid carcinoma, the patient’s age (FIG-5) plays a crucial role in the stage of the disease.

**Fig. 5:**
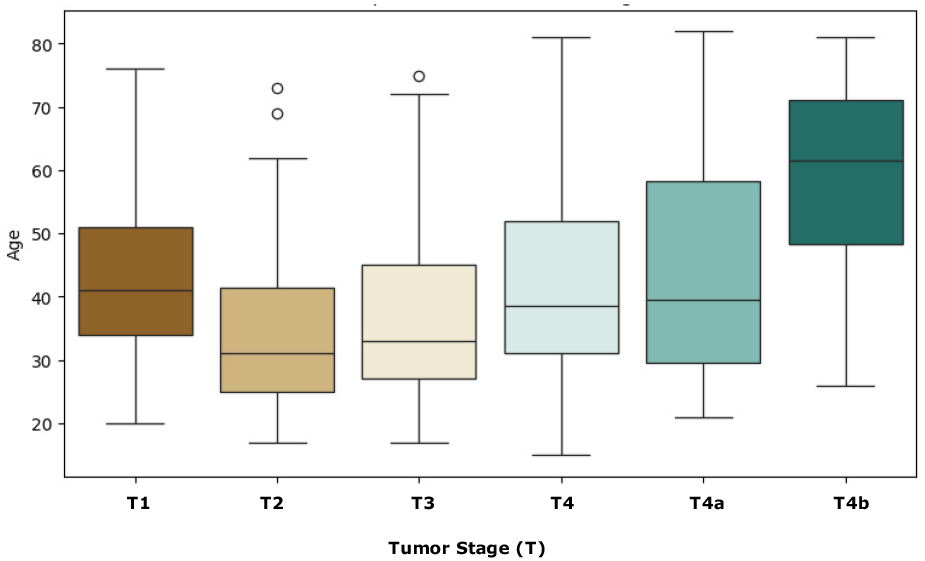
Age distribution based on different stages of Tumor

By analyzing pathological reports, it has been observed that thyroid cancer is significantly more common in women than in men. Women are approximately three times more likely to develop thyroid cancer. The reasons for this disparity are thought to be related to hormonal factors, especially estrogen. Figure-6 describes that women are at a higher risk of developing all types of thyroid cancer (Follicular, Papillary, Micropapillary, Hurthel cell).

**Fig. 6:**
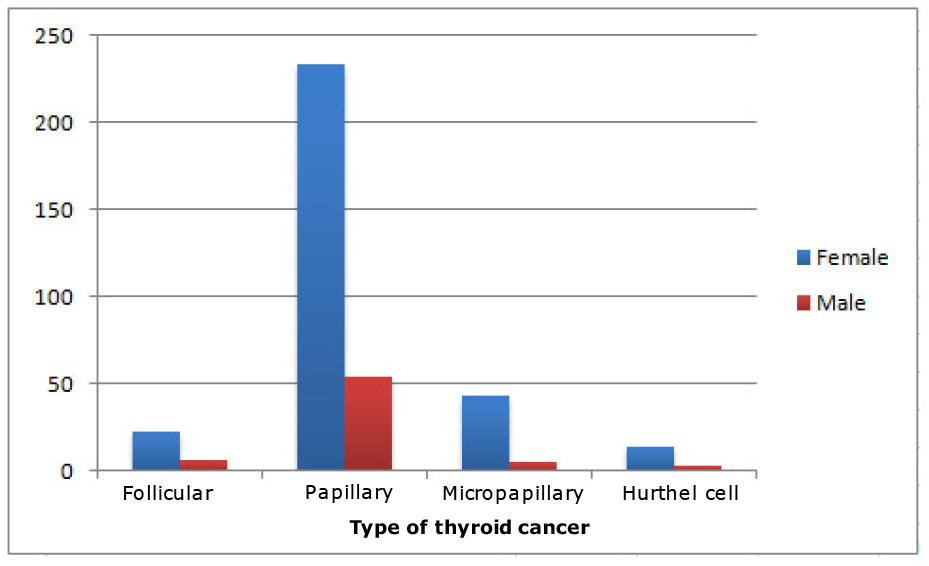
Gender-Based Risk Analysis of Thyroid Cancer

#### B. Feature extraction from Ultrasound Image Analysis

In this study, we have also considered different features that are extracted from ultrasound image analysis. To perform a comprehensive thyroid ultrasound image analysis for cancer detection, we need to segment the thyroid region using edge detection and contour detection techniques. Several features that are relevant to cancer detection are extracted from the ultrasound image.

#### 1) Nodule Detection and Composition Analysis

After segmentation, largest contour has been chosen (Fig-7) to identify the correct nodule, and location of the nodule has been extracted by computing the bounding box of the contour to get the location coordinates. We have also identify the peripheral halo, which is a hypoechoic (darker) ring surrounding a nodule, often indicating a benign nodule.

**Fig. 7:**
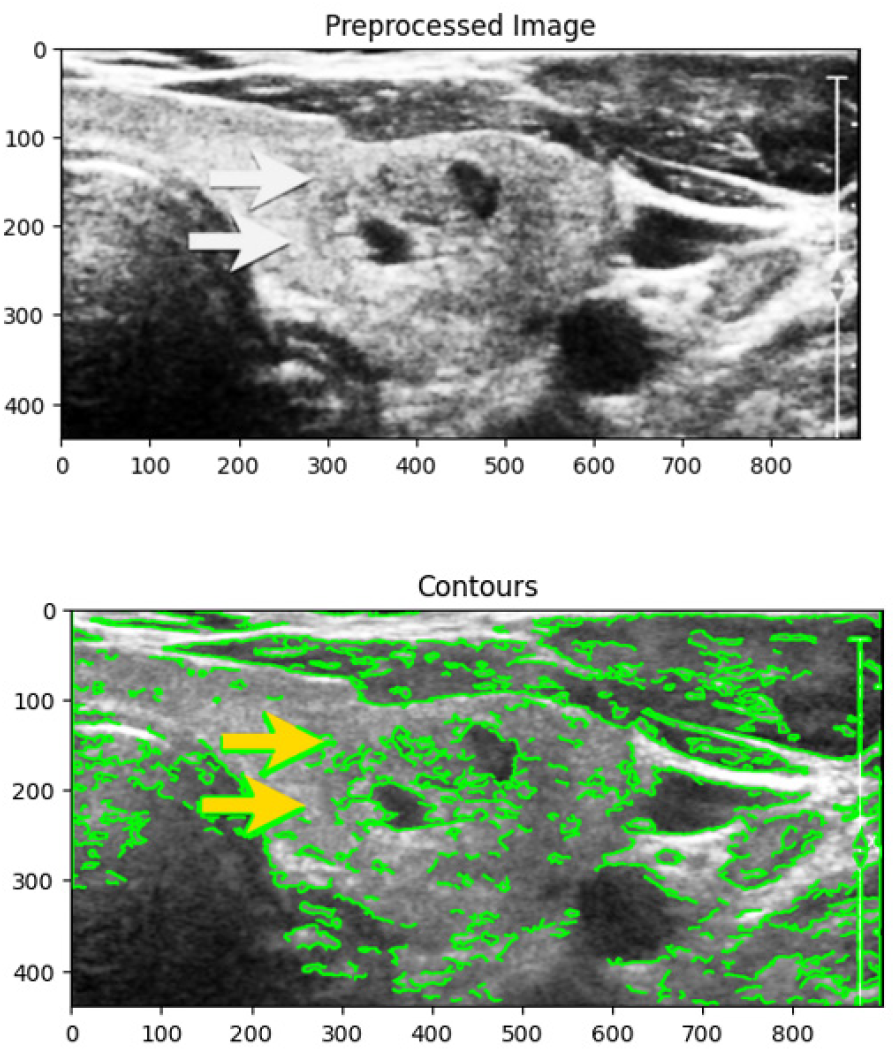
Segmentation of the thyroid region using edge detection and contour detection techniques.

Composition analysis (Fig-8) of the identified nodule is performed to calculate the mean intensity of the nodule region and determine the proportion of pixels below (cystic) and above (solid) certain intensity thresholds. The nodule is classified based on these proportions into different categories: cystic, solid, predominantly cystic, predominantly solid, or complex.

**Fig. 8:**
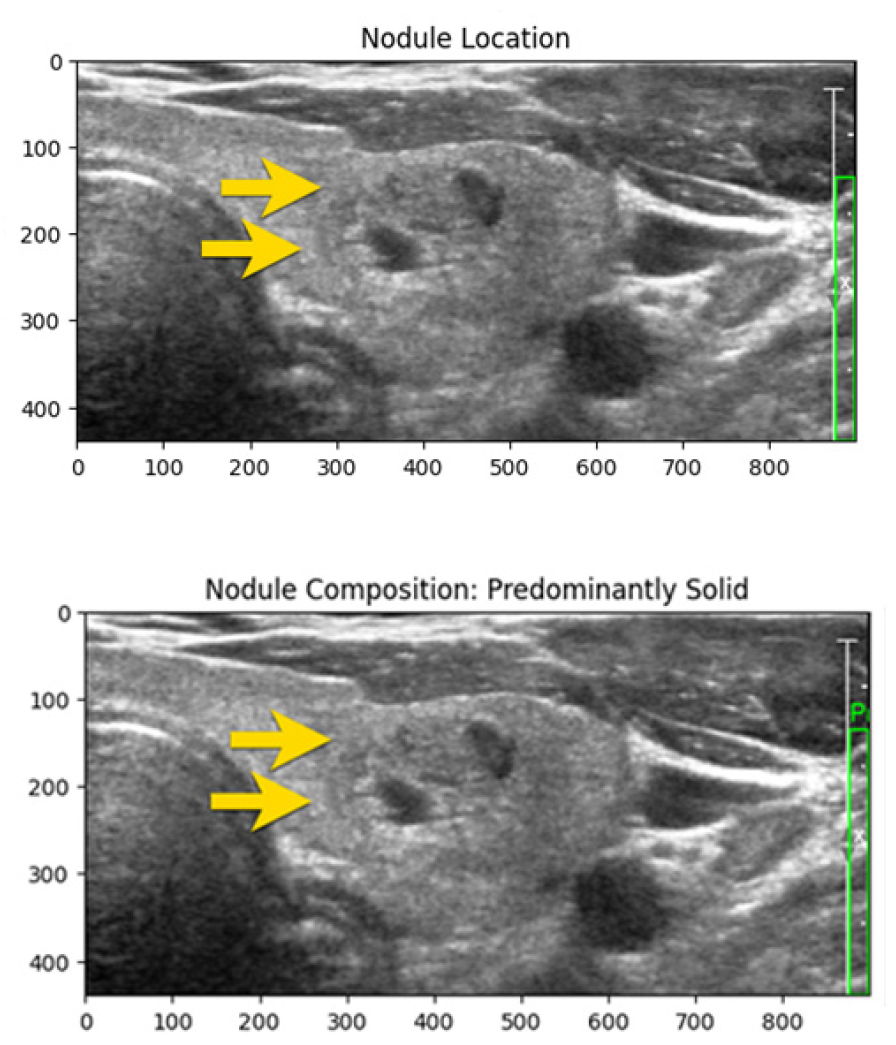
Nodule Detection and Composition Analysis

#### 2) Vascularity Analysis Of Nodule

Vascularity analysis (Fig-9) in thyroid ultrasound images is important for evaluating the blood flow in and around the thyroid nodule. Increased vascularity can be a marker for malignancy. Doppler ultrasound is commonly used to assess vascularity by detecting and visualizing blood flow.

**Fig. 9:**
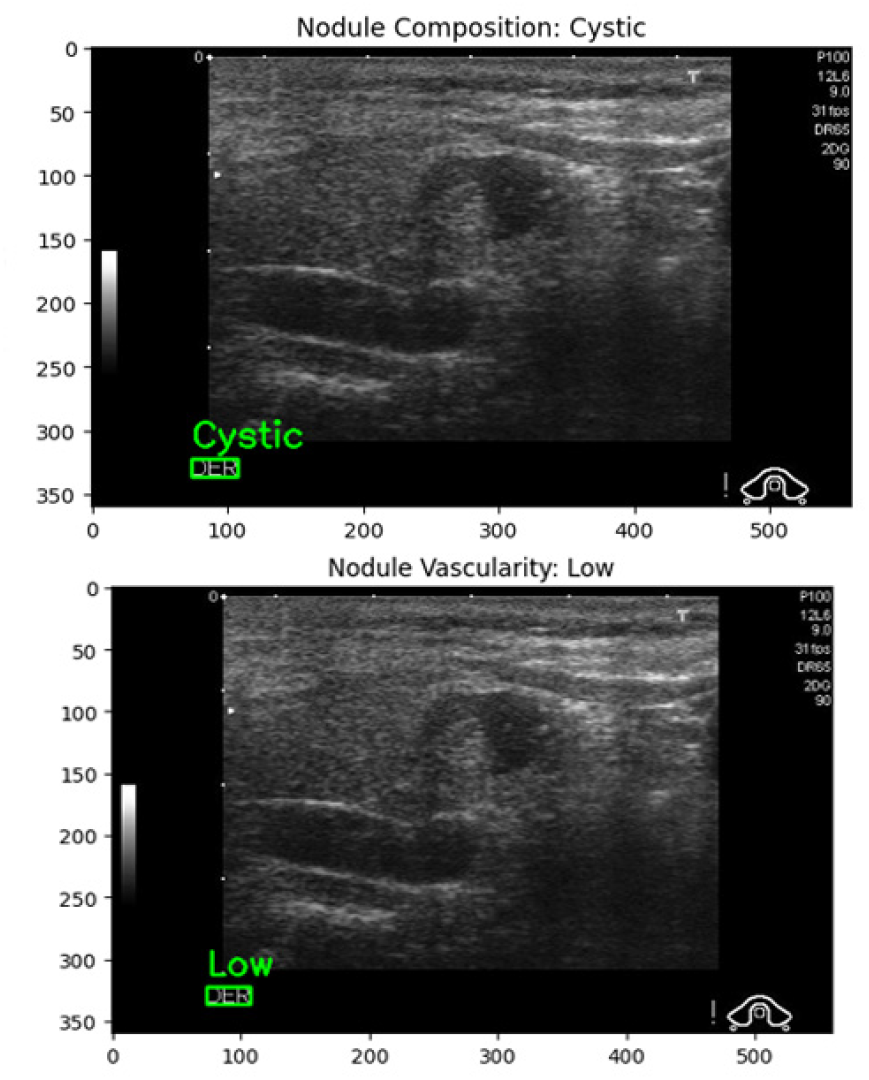
Vascularity Analysis Of Nodule

Echogenic foci (Fig-10) are bright spots in ultrasound images, often indicating microcalcifications within thyroid nodules. The presence of echogenic foci can be a significant marker for malignancy. Table-II described few important features those are extracted from different ultrasound samples.

**Fig. 10:**
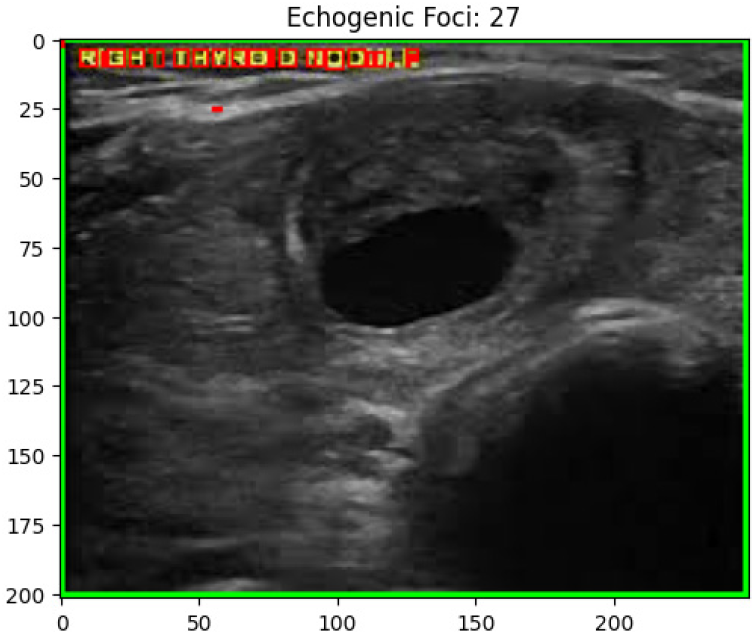
Echogenic foci Detection

**TABLE II:** Few Important Ultrasonographic Features (Nodule)

| Sample | Size (mm) | Shape | Margins | Vascularity | Echogenicity | Calcification | Composition | Echogenic Foci |
| --- | --- | --- | --- | --- | --- | --- | --- | --- |
| S1 | 34 | Irregular | Irregular | Low | Hypoechoic | Microcalcifications | Cystic | 3 |
| S2 | 256 | Irregular | Irregular | Low | Isoechoic | Microcalcifications | Pre-dominantly Cystic | 12 |
| S3 | 80 | round/oval | Irregular | Low | Isoechoic | Macrocalcifications | Pre-dominantly Solid | 2 |
| S4 | 26 | Irregular | Irregular | Low | Hyperechoic | Macrocalcifications | Pre-dominantly Solid | 2 |
| S5 | 248 | Irregular | Irregular | Low | Isoechoic | Microcalcifications | Cystic | 27 |
| S6 | 32 | Irregular | Irregular | Low | Isoechoic | Macrocalcifications | Solid | 1 |

#### C. Clinical notes Analysis

Medical terms are extracted from clinical notes using the BioClinicalBERT model from the EHR (Electronic Health Record System), transformed into features, integrated with existing structured data, and utilized to train a machine learning model for disease detection.

After extracting features from both the clinical text and the scanned images, these are passed through Cross-Modal Attention Fusion (Table-III).

**TABLE III:** Performance of Cross-Modal Attention Fusion for Thyroid Cancer Prediction.

| Model | Accuracy (%) | Precision (%) | Recall (%) | F1-score (%) |
| --- | --- | --- | --- | --- |
| ViT (MRI Only) | 88.6 | 87.9 | 86.8 | 87.3 |
| BioClinicalBERT (Text Only) | 85.2 | 84.5 | 83.7 | 84.1 |
| Early Feature Fusion | 90.4 | 89.7 | 88.9 | 89.3 |
| Cross-Modal Attention Fusion | **94.1** | **93.5** | **92.8** | **93.1** |

**TABLE IV:**
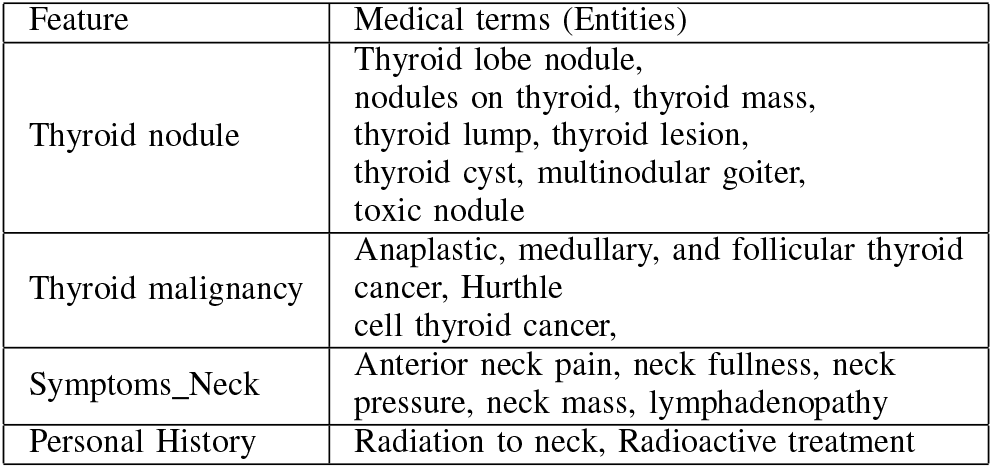
Extracted features from clinical notes.

| Feature | Medical terms (Entities) |
| --- | --- |
| Thyroid nodule | Thyroid lobe nodule, nodules on thyroid, thyroid mass, thyroid lump, thyroid lesion, thyroid cyst, multinodular goiter, toxic nodule |
| Thyroid malignancy | Anaplastic, medullary, and follicular thyroid cancer, Hurthle cell thyroid cancer, |
| Symptoms_Neck | Anterior neck pain, neck fullness, neck pressure, neck mass, lymphadenopathy |
| Personal History | Radiation to neck, Radioactive treatment |

**TABLE V:**
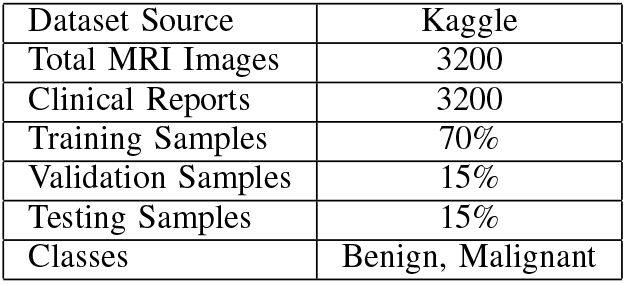
Dataset Description.

#### D. Train and evaluate a machine learning model using the multimodal dataset

The multimodal framework trains on publicly available thyroid cancer datasets (Table-**??** which researchers have collected from the Kaggle platform. The dataset contains MRI images together with linked clinical text data, which includes medical reports and prescription notes.

The experimental results demonstrate that the proposed multimodal framework achieves superior performance compared to state-of-the-art models. The Vision Transformer effectively captures spatial features from MRI images, while BioClinical-BERT extracts contextual medical information from clinical text. The cross-modal attention fusion mechanism enables the model to learn interactions between visual and textual representations, leading to improved diagnostic performance. The proposed model achieves 94.3% accuracy which represents the highest accuracy according to Table VI while surpassing the performance of unidirectional models which include CNN and ResNet and standalone ViT models. The evidence shows that multimodal data integration improves both reliability and accuracy in predicting thyroid cancer. The graphical representation in Fig. 11 shows that the proposed multimodal framework significantly outperforms the unimodal baseline models across all evaluation metrics. By integrating MRI imaging features with contextual clinical text information, the model achieves improved accuracy, sensitivity, specificity, and AUC, demonstrating its effectiveness for reliable thyroid cancer prediction and supporting better preoperative clinical decision-making.

**TABLE VI:** Accuracy Comparison with State-of-the-Art Models.

| Model | Modality | Accuracy (%) |
| --- | --- | --- |
| CNN [25] | MRI Images | 84.4 |
| ResNet-50 [27] | MRI Images | 85.7 |
| Vision Transformer (ViT) [28] | MRI Images | 89.2 |
| BERT-based Model[30] | Clinical Text | 87.6 |
| BioClinicalBERT [26] | Clinical Text | 85.1 |
| Multimodal CNN + RNN | Image [26]+ Text | 89.5 |
| **Proposed Model(ViT + BioClinicalBERT (Cross-modal Fusion))** | Image + Text | **92.5** |

**Fig. 11:**
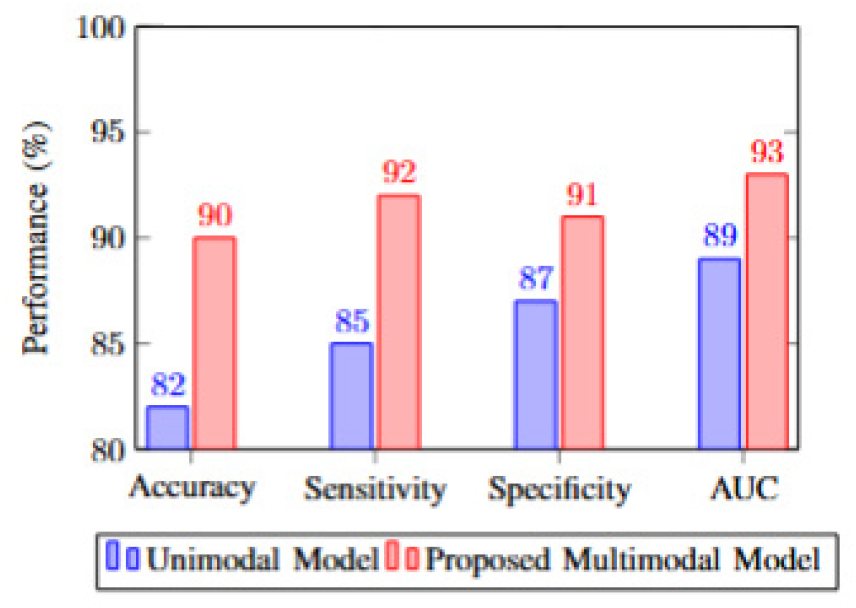
Performance comparison between the unimodal and proposed multimodal system.

## V. Conclusion and Future Work

This study developed a deep learning framework that uses multimodal input to predict thyroid cancer by combining MRI scanned image with clinical text information. The Vision Transformer (ViT) was employed to extract spatial features from MRI scans, while BioClinicalBERT was used to capture contextual information from clinical reports and prescriptions. The research introduced a cross-modal attention fusion mechanism, which helps the model to comprehend important connections between its visual content and textual information. The experimental results with the Kaggle dataset showed that the proposed method outperformed multiple state-of-the-art systems, which achieved greater accuracy in classifying both benign and malignant thyroid tumors. The results demonstrate that multimodal learning provides better diagnostic accuracy while eliminating the restrictions that single-modality systems face.

The proposed framework provides support to clinicians who need to detect early thyroid cancer while they make medical decisions. In future work, the model can be extended by incorporating additional medical modalities such as ultrasound images, genomic data, and larger clinical datasets to further enhance prediction performance and generalization.

## a) Funding

Not Applicable

## b) Ethical Approval

Not Applicable

## c) Informed Consent

This article does not contain any studies with human participants performed by any of the authors.

## d) Authors Contribution Statement

The authors confirm contribution to the paper as follows: study conception, analysis and interpretation of results by Paramita Ray. Paramita Ray is also involved in manuscript preparation.

## e) Competing Interests

The author whose name is listed above certifies that she has nO affiliations with or involvement in any organization or entity with any financial interest (such as honorarium; educational grants; participation in speakers bureaus; membership, employment, consultancies, stock ownership, or other equity interest; and expert testimony or patentlicensing arrangements) or non-financial interest (such as personal or professional relationships, affiliations, knowledge or beliefs) in the subject matter or materials discussed in this manuscript.

## f) Data Availability Statement

All data analyzed or generated are included in the paper. **Source Data:** Data sets are collected from Kaggle. (https://www.kaggle.com/datasets/ankushpanday1/thyroid-cancer-risk-prediction-dataset)

## Notes

### Competing Interest Statement

The authors have declared no competing interest.

## References

[1] G. Litjens et al., “A survey on deep learning in medical image analysis,” Medical Image Analysis, vol. 42, pp. 60–88, 2017.

[2] A. Dosovitskiy et al., “An image is worth 16×16 words: Transformers for image recognition at scale,” in Proc. Int. Conf. Learning Representations (ICLR), 2021.

[3] H. Li et al., “Computer-aided diagnosis of thyroid nodules using deep learning on ultrasound images,” IEEE Transactions on Medical Imaging, vol. 38, no. 4, pp. 901–910, 2019.

[4] S. Y. Ko et al., “Application of deep learning to thyroid ultrasound imaging for cancer detection,” Scientific Reports, vol. 10, 2020.

[5] J. Devlin, M.-W. Chang, K. Lee, and K. Toutanova, “BERT: Pre-training of deep bidirectional transformers for language understanding,” in Proc. NAACL-HLT, 2019, pp. 4171–4186.

[6] E. Alsentzer et al., “Publicly available clinical BERT embeddings,” in Proc. 2nd Clinical Natural Language Processing Workshop, 2019, pp. 72–78.

[7] K. Singhal et al., “Large language models encode clinical knowledge,” Nature, vol. 620, pp. 172–180, 2023.

[8] G. Huang et al., “Fusion of medical imaging and electronic health records using deep learning: A review,” Medical Image Analysis, vol. 67, 2020.

[9] D. Zhang et al., “Multimodal deep learning for healthcare applications: A review,” IEEE Journal of Biomedical and Health Informatics, vol. 25, no. 9, pp. 3503–3517, 2021.

[10] A. Radford et al., “Learning transferable visual models from natural language supervision,” in Proc. Int. Conf. Machine Learning (ICML), 2021.

[11] Y. Gu et al., “Domain-specific language model pretraining for biomedical natural language processing,” ACM Transactions on Computing for Healthcare, vol. 3, no. 1, pp. 1–23, 2021.

[12] I. Beltagy, K. Lo, and A. Cohan, “SciBERT: A pretrained language model for scientific text,” in Proc. Conf. Empirical Methods in Natural Language Processing (EMNLP), 2019, pp. 3615–3620.

[13] K. Simonyan and A. Zisserman, “Very deep convolutional networks for large-scale image recognition,” in Proc. Int. Conf. Learning Representations (ICLR), 2015.

[14] K. He, X. Zhang, S. Ren, and J. Sun, “Deep residual learning for image recognition,” in Proc. IEEE Conf. Computer Vision and Pattern Recognition (CVPR), 2016, pp. 770–778.

[15] N. Srivastava and R. Salakhutdinov, “Multimodal learning with deep Boltzmann machines,” Journal of Machine Learning Research, vol. 15, pp. 2949–2980, 2014.

[16] E. Alsentzer et al., “Publicly available clinical BERT embeddings,” in Proc. Clinical Natural Language Processing Workshop, 2019.

[17] K. Singhal et al., “Large language models encode clinical knowledge,” Nature, vol. 620, pp. 172–180, 2023.

[18] G. Huang et al., “Fusion of medical imaging and electronic health records using deep learning: A review,” Medical Image Analysis, vol. 67, 2020.

[19] D. Zhang et al., “Multimodal deep learning for healthcare applications: A review,” IEEE Journal of Biomedical and Health Informatics, vol. 25, no. 9, pp. 3503–3517, 2021.

[20] A. Radford et al., “Learning transferable visual models from natural language supervision,” in Proc. Int. Conf. Machine Learning (ICML), 2021.

[21] A. Dosovitskiy et al., “An image is worth 16×16 words: Transformers for image recognition at scale,” in Proc. Int. Conf. Learning Representations (ICLR), 2021.

[22] E. Alsentzer, J. Murphy, W. Boag, W.-H. Weng, D. Jin, T. Naumann, and M. McDermott, “Publicly available clinical BERT embeddings,” in Proc. 2nd Clinical Natural Language Processing Workshop, Minneapolis, MN, USA, 2019, pp. 72–78.

[23] J. Devlin, M.-W. Chang, K. Lee, and K. Toutanova, “BERT: Pre-training of deep bidirectional transformers for language understanding,” in Proc. NAACL-HLT, Minneapolis, MN, USA, 2019, pp. 4171–4186.

[24] Y. Gu et al., “Domain-specific language model pretraining for biomedical natural language processing,” ACM Transactions on Computing for Healthcare, vol. 3, no. 1, pp. 1–23, 2021.

[25] MathWorks, “Train vision transformer network for image classification,” [Online]. Available: https://www.mathworks.com/help/deeplearning/ug/train-vision-transformer-network-for-image-classification.html

[26] K. Simonyan and A. Zisserman, “Very deep convolutional networks for large-scale image recognition,” in Proc. Int. Conf. Learning Representations (ICLR), 2015.

[27] K. He, X. Zhang, S. Ren, and J. Sun, “Deep residual learning for image recognition,” in Proc. IEEE Conf. Computer Vision and Pattern Recognition (CVPR), 2016, pp. 770–778.

[28] J. Devlin, M. Chang, K. Lee, and K. Toutanova, “BERT: Pre-training of deep bidirectional transformers for language understanding,” in Proc. NAACL-HLT, 2019, pp. 4171–4186.

[29] N. Srivastava and R. Salakhutdinov, “Multimodal learning with deep Boltzmann machines,” Journal of Machine Learning Research, vol. 15, pp. 2949–2980, 2014.

[30] E. Alsentzer, J. Murphy, W. Boag, W.-H. Weng, D. Jin, T. Naumann, and M. McDermott, “Publicly available clinical BERT embeddings,” in Proc. 2nd Clinical Natural Language Processing Workshop, Minneapolis, MN, USA, 2019, pp. 72–78.

